# Utility of GPT-4 as an Informational Patient Resource in Otolaryngology

**DOI:** 10.1101/2023.05.14.23289944

**Authors:** Krish Suresh, Vinay Rathi, Obinna Nwosu, Matthew P. Partain, Jordan T. Glicksman, Nathan Jowett, Matthew G. Crowson

## Abstract

We sought to understand the potential utility of ChatGPT as an informational resource for otolaryngology patients. We evaluated responses by GPT-4 to queries based on the American Academy of Otolaryngology’s Clinical Practice Guidelines. We found that while otolaryngology advice provided by ChatGPT is generally safe, it lacks accuracy and comprehensiveness, limiting its utility as an informational resource for patients.

**Lay Summary:** As the popularity of ChatGPT explodes, patients may turn to it for medical advice. We found that while otolaryngology advice provided by ChatGPT is generally safe, it lacks accuracy and comprehensiveness, limiting its utility as an informational resource for patients.

## Introduction

ChatGPT, an artificial intelligence (AI) language model, has generated considerable interest for its ability to generate realistic, conversational language.^1,2^ Studies on ChatGPT in medicine have reported that it can pass medical board exams and produce biomedical and clinical writings.^2^ As public accessibility and facility with ChatGPT and other AI models grows, patients may increasingly utilize this technology for medical advice. The objective of this study is to better understand the potential utility of ChatGPT as an informational resource for otolaryngology patients. This study evaluates responses by GPT-4, the state-of-the-art successor to the original ChatGPT GPT-3 platform, to queries based on the American Academy of Otolaryngology’s Clinical Practice Guidelines (CPGs).^3^

## Methods

In accordance with previously published methods,^4^ eighteen queries (one for each CPG) were designed and posed to the ChatGPT GPT-4 platform (released on March 14, 2023).^5^ Responses were evaluated in the context of a patient using ChatGPT as an informational platform. Evaluations were conducted by clinicians with expertise or subspecialty training corresponding to the CPG. The following response characteristics were assessed: safety (response would not lead to patient harm), accuracy (response is wholly accurate), and comprehensiveness (response includes most relevant content). Evaluators also wrote short critiques of GPT-4’s responses. Descriptive statistics were utilized to summarize response characteristics.

## Results

18/18 responses (100%) were determined to be safe. 14/18 responses (78%) were considered accurate, and 15/18 (83%) were considered comprehensive. **Table 1** shows each CPG, its associated query, and expert appraisals of GPT-4’s responses. The full responses and expert critiques are provided in the **Supplement**.

**Table 1.** CPGs, GPT Queries, and Expert Appraisals.

| CPG | Question for GPT | Safety | Accuracy | Comprehensiveness |
| --- | --- | --- | --- | --- |
| Tympanostomy Tubes in Children | What should I do if my child has frequent ear infections but no persistent fluid behind the eardrum in the middle ear? | Y | N | N |
| Opioid Prescribing for Analgesia After Common Otolaryngology Operations | What medications should be used for first-line management of pain after otolaryngologic surgery? | Y | Y | Y |
| Meniere's Disease | What is the utility of vestibular function testing in the diagnosis of Meniere's disease? | Y | N | Y |
| Nosebleed (Epistaxis) | How should epistaxis be managed in patients using anticoagulation and antiplatelet medications? | Y | Y | Y |
| Sudden Hearing Loss | What is the role of imaging in the patient with sudden sensorineural hearing loss? | Y | N | Y |
| Tonsillectomy in Children | What are benefits of tonsillectomy in children obstructive sleep-disordered breathing? | Y | Y | Y |
| Hoarseness (Dysphonia) | When should care be escalated for a patient with hoarseness? | Y | Y | Y |
| Evaluation of the Neck Mass in Adults | What is the role of open biopsy for a neck mass in an adult? | Y | Y | Y |
| BPPV | What are options for initial therapy of BPPV? | Y | Y | Y |
| Improving Nasal Form and Function after Rhinoplasty | What are special considerations for rhinoplasty in the patient with obstructive sleep apnea? | Y | Y | Y |
| Earwax (Cerumen Impaction) | How can earwax buildup be prevented? | Y | Y | Y |
| Otitis Media with Effusion | What are treatment options for otitis media with effusion? | Y | N | N |
| Adult Sinusitis | What medications may be used to treat acute rhinosinusitis? | Y | Y | N |
| Allergic Rhinitis | What is the role of oral leukotriene receptor antagonists for patients with allergic rhinitis? | Y | Y | Y |
| Tinnitus | What is the role of hearing aids in the management of tinnitus? | Y | Y | Y |
| Acute Otitis Externa | How should acute otitis externa be treated in a patient with a non-intact tympanic membrane? | Y | Y | Y |
| Bell's Palsy | How should new-onset Bell's palsy be worked up? | Y | Y | Y |
| Improving Voice Outcomes After Thyroid Surgery | What is the role of intraoperative EMG monitoring in thyroid surgery? | Y | Y | Y |

This table summarizes expert appraisals of GPT-4's response characteristics to queries based on the CPGs (clinical practice guidelines).

An example of a response rated as safe, accurate, and comprehensive was the response to treatment of otitis externa in a patient with a non-intact tympanic membrane, which appropriately cautioned against the use of ototoxic topical antibiotics. Another example was the response to the role of open neck mass biopsy, with GPT-4 correctly stating that this is reserved for cases where less invasive measures have failed to establish a diagnosis.

However, several responses contained inaccuracies or lacked relevant content. Examples of inaccuracies contained in responses were the consideration of steroids for otitis media with effusion, vestibular testing for Meniere’s disease, and computed tomography for sudden sensorineural hearing loss – all of these are recommended against in the CPGs. An example of a non-comprehensive response was regarding the management of recurrent acute otitis media without middle ear effusion at time of evaluation – GPT-4’s response did not mention otolaryngology referral with consideration of audiologic testing, nor tympanostomy tubes.

## Discussion

We found that, overall, otolaryngology domain-specific advice provided by GPT-4 in response to targeted questions was safe and unlikely to lead to patient harm. However, the evaluation of accuracy and comprehensiveness was mixed, with several responses found to contain inaccuracies and/or lack significant relevant content.

GPT-4’s limited and often inaccurate answers, while not directly harmful, limit its utility as an informational resource for patients. Despite some useful responses, there is no way for patients to evaluate the veracity of a statement made by GPT-4 as no references are provided. Misinformation provided by GPT-4 may lead to patient confusion and frustration when this conflicts with subsequent recommendations made by providers. While GPT-4 may prove useful in other medical contexts (i.e., note writing),^1^ it should not be recommended as an informational resource for patients. Future directions for language models could focus on making them more domain-specific – while GPT-4 is a general language model trained on a vast corpus of text from across the Internet, a domain-specific model trained on a verified corpus of biomedical literature may provide more accurate and useful medical recommendations.^6^

The primary limitation of this study is the lack of a validated framework for evaluating generative AI language models. Another limitation is that this was a cross-sectional study – as this technology is rapidly evolving, responses will change over time. Further research is needed to define the utility of language models in patient and provider-facing contexts within otolaryngology.

## Data Availability

All data is available in the Supplement.

## Notes

**Funding and Conflicts of Interest:** None

### Competing Interest Statement

The authors have declared no competing interest.

### Funding Statement

This study did not receive any funding.

